# Longitudinal Validation of a Deep Learning Index for Aortic Stenosis Progression

**DOI:** 10.1101/2025.02.17.25322392

**Authors:** Jiesuck Park, Jiyeon Kim, Yeonyee E. Yoon, Jaeik Jeon, Seung-Ah Lee, Hong-Mi Choi, In-Chang Hwang, Goo-Yeong Cho, Hyuk-Jae Chang, Jae-Hyeong Park

**Affiliations:** Cardiovascular Center and Division of Cardiology, Department of Internal Medicine, Seoul National University Bundang Hospital, Seongnam, Gyeonggi, Republic of Korea; Department of Internal Medicine, Seoul National University College of Medicine, Seoul, Republic of Korea; Ontact Health Inc., Seoul, Republic of Korea; CONNECT-AI Research Center, Yonsei University College of Medicine, Seoul, Republic of Korea; Division of Cardiology, Severance Cardiovascular Hospital, Yonsei University College of Medicine, Yonsei University Health System, Seoul, Republic of Korea; Department of Cardiology, Internal Medicine, Chungnam National University Hospital, 282 Munhwa-ro, Jung-gu Daejeon, 35015, Republic of Korea

**Keywords:** Aortic stenosis, artificial intelligence, echocardiography, progression

## Abstract

**Background:** Aortic stenosis (AS) is a progressive disease requiring timely monitoring and intervention. While transthoracic echocardiography (TTE) remains the diagnostic standard, deep learning (DL)-based approaches offer potential for improved disease tracking. This study examined the longitudinal changes in a previously developed DL-derived index for AS continuum (DLi-ASc) and assessed its value in predicting progression to severe AS.

**Methods:** We retrospectively analysed 2,373 patients (7,371 TTEs) from two tertiary hospitals. DLi-ASc (scaled 0-100), derived from parasternal long- and/or short-axis views, was tracked longitudinally. The median follow-up duration was 42.8 months (IQR 22.2–75.7 months).

**Results:** DLi-ASc increased in parallel with worsening AS stages (p for trend <0.001) and showed strong correlations with AV maximal velocity (V_max_) (Pearson correlation coefficients [PCC] = 0.69, p<0.001) and mean pressure gradient (mPG) (PCC = 0.66, p<0.001). Higher baseline DLi-ASc was associated with a faster AS progression rate (p for trend <0.001). Additionally, the annualized change in DLi-ASc, estimated using linear mixed-effect models, correlated strongly with the annualized progression of AV V_max_ (PCC = 0.71, p<0.001) and mPG (PCC = 0.68, p<0.001). In Fine-Gray competing risk models, baseline DLi-ASc independently predicted progression to severe AS, even after adjustment for AV V_max_ or mPG (hazard ratio per 10-point increase = 2.38 and 2.80, respectively)

**Conclusion:** DLi-ASc increased in parallel with AS progression and independently predicted severe AS progression. These findings support its role as a non-invasive imaging-based digital marker for longitudinal AS monitoring and risk stratification.

**CLINICAL PERSPECTIVE:** *What Is New?:*

- This is the first study to validate longitudinal changes in a deep learning-derived index (DLi-ASc) for tracking aortic stenosis (AS) progression.
- DLi-ASc increases consistently over time in parallel with worsening AS stages and conventional AS hemodynamic parameters.
- Baseline DLi-ASc independently predicts future severe AS progression, even after adjusting for conventional hemodynamic parameters.

*What Are the Clinical Implications?:*

- DLi-ASc provides a quantitative, noninvasive digital marker for monitoring AS progression in routine clinical practice.
- DLi-ASc enables individualized risk stratification and may inform tailored follow-up strategies for patients with AS.
- DLi-ASc may serve as a surrogate marker for future studies evaluating therapeutic interventions to slow AS progression.

## 1. INTRODUCTION

Aortic stenosis (AS) is a progressive disease that worsens over time and is increasingly prevalent in aging populations.^1,2^ Effective management requires timely monitoring and intervention. Transthoracic echocardiography (TTE) remains the gold standard for AS evaluation,^3,4^ but its reliance on multiple Doppler-derived parameters introduces variability and requires operator expertise for accurate image acquisition and interpretation. To enhance objectivity and scalability, artificial intelligence (AI) has been explored for AS assessment, primarily in two directions: (1) automating conventional AS evaluation and (2) predicting significant or severe AS using limited imaging data.^5–8^

A number of deep learning (DL)-based indices for AS severity have recently been developed,^5–7^ each differing in input views, labelling strategies, and modelling approaches. Among these, we developed a novel DL-derived index for the AS continuum (DLi-ASc) that can quantify AS severity using limited TTE views – either parasternal long-axis (PLAX) or parasternal short-axis (PSAX) views – within a multi-task learning framework, where an ordinal AS severity grade prediction was combined with conventional echocardiographic parameter prediction to produce a continuous AS severity index, thereby enhancing robustness and interpretability.^5^ Rigorous validation demonstrated its diagnostic accuracy and prognostic value, comparable to conventional AS parameters.

Some of these indices have also been evaluated for prognostic value in longitudinal follow-up.^9,10^ However, prior longitudinal studies have predominantly relied on baseline DL scores to predict progression or outcomes, without systemically characterizing temporal score trajectories or establishing their relationship to conventional hemodynamic progression or clinical outcomes.

In the present study, we extended the application of DLi-ASc to longitudinal AS assessment, aiming to: (1) characterize temporal changes in DLi-ASc and express them as annualized rates; (2) examine whether these changes align with the progression of conventional echocardiographic parameters; and (3) evaluate the prognostic value of both baseline DLi-ASc and its annualized rate of change for severe AS progression. By addressing these gaps, this study aims to provide one of the first demonstrations that a DL-derived AS index can capture disease trajectory over time, offering a standardized and interpretable marker with potential value for risk prediction and long-term monitoring.

## 2. METHODS

The data that support the findings of this study are available upon reasonable request. Please contact the corresponding author to request the minimal anonymized dataset. Researchers with additional inquiries about the deep learning model developed in this study are also encouraged to reach out to the corresponding author.

### 2.1. Study Population

The study population was collected from two tertiary institutions in South Korea: Seoul National University Bundang Hospital (SNUBH, March 2006 to October 2024) and Chungnam National University Hospital (CNUH, February 2010 to October 2024). Patients who underwent at least two serial TTE examinations and were diagnosed with AS, either at the time of the initial TTE or during follow-up, were included. A total of 2,806 patients with 8,663 TTE examinations were initially identified (SNUBH: 1,639 patients, 5,049 TTEs; CNUH: 1,167 patients, 3,614 TTEs) To ensure the methodological consistency and reliability of AS severity assessment, the following exclusion criteria were applied based on baseline TTE findings: 1) severe AS, 2) ≥ moderate aortic regurgitation, 3) ≥ moderate mitral stenosis or regurgitation, 4) discordant AS classification, defined as cases where aortic valve (AV) maximal velocity (V_max_), mean pressure gradient (mPG), and AV area (AVA) fell into different severity categories (**Figure 1**). After applying these exclusion criteria, 2,373 patients with 7,371 TTE examinations remained for analysis. This study was approved by the institutional review boards of the participating hospitals, with a waiver for informed consent due to its retrospective, observational design (SNUBH B-2302-808-101; CNUH 2024-11-019). The study adhered to the revision of the Declaration of Helsinki (2013).

**Figure 1.**
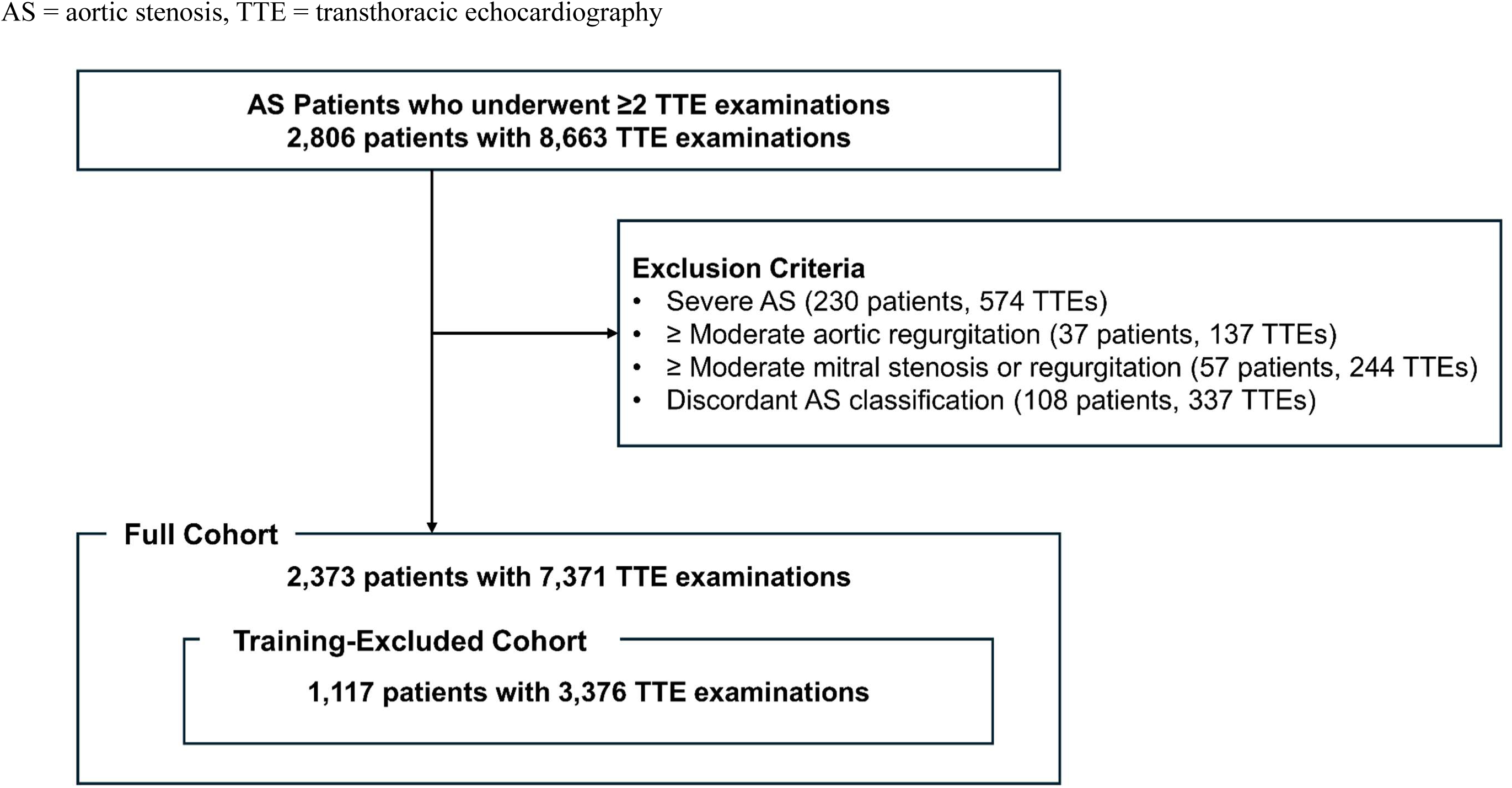
Study Population.

### 2.2. Clinical and Echocardiographic Data Acquisition

All TTE examinations were performed by trained echocardiographers or cardiologists following contemporary guidelines^3,4^ and interpreted by board-certified cardiologists who specialized in echocardiography. AV sclerosis was defined as focal or diffuse thickening and calcification with V_max_ <2.0 m/sec and preserved leaflet motion. For follow-up TTEs, when parameter discordance between Vmax, mPG, and AVA was observed, AS severity was determined based on the final interpretation documented by the attending cardiologist. In cases with indeterminate severity or potential low-flow states, if additional studies, such as dobutamine stress TTE or CT-based AV calcium scoring, had been performed, the results were reviewed and incorporated into the final clinical interpretation. All available TTE examinations for each patient were collected, for those who underwent AV replacement during the follow-up period, only TTE data obtained before the procedure were included. AS severity metrics from each TTE were extracted from clinical reports used for the treatment decision-making.^3^

### 2.3. DL-Based AS Assessment Algorithm

The DL model used in this study has been previously described and is integrated into an AI-enhanced AS evaluation system (USfeat_valve.ai, Ontact Health, Korea).^5^ Briefly, the model utilizes a 3-dimensional (3D) convolutional neural network (r2plus1d18) to analyse TTE videos, generating a continuous severity score, the DLi-ASc (range 0–100). This index quantifies AS severity based on limited TTE views - specifically the PLAX and PSAX views at the aortic valve (AV) level. To enhance the reliability of AS severity evaluation, the model incorporates ordered label mapping and multi-task learning, leveraging auxiliary tasks to predict key TTE parameters such as AV maximal velocity (V_max_), mean pressure gradient (mPG), and AV area (AVA). For each patient, if multiple PLAX or PSAX views were available, DLi-ASc was first averaged within each view type (PLAX and PSAX separately) and then averaged across both views. If only one view type was available, its score was used. The DLi-ASc was generated for all baseline and follow-up TTE examinations included in this study. The DL model successfully generated DLi-ASc for all cases, ensuring full data availability for longitudinal analysis with no exclusions due to model processing failure.

### 2.4. Statistical Analysis

All statistical analyses were conducted in both the full cohort and the training-excluded cohort, which excluded all patients whose TTE examinations were used in the training phase of the DLi-ASc model (**Figure 1**).^5^ This parallel analytical approach was implemented to ensure the robustness and generalizability of our findings, while addressing concerns regarding potential data leakage and overfitting.^11^

Clinical data are described in the study as the median with interquartile range (IQR) or mean with 95% confidence interval (CI) for continuous variables and as percentage numbers for factorial variables. Baseline characteristics were compared between the cohorts, with significant differences determined using the Kruskal-Wallis test and Chi-square test, as appropriate.

The temporal progression of AS severity was illustrated using a proportional stacked bar plot, showing the relative distribution of AS stage (normal AV, AV sclerosis, mild AS, moderate AS, and severe AS) over the follow-up period. This trend was further stratified according to baseline AS severity categories. To assess the significance of ordinal changes in AS stage over time, we employed a Cumulative Link Mixed Model, which accounts for repeated measurements and varying intervals between TTEs.^12^ The temporal changes of DLi-ASc were visualized using ridge density plots and analysed using a Linear Mixed Model (LMM), adjusting for repeated TTE measurements and irregular follow-up intervals.^13^ The same modelling approach was applied to AV V_max_ and AV mPG, enabling estimation of annual progression rates based on all available serial measurements.

The relationships between DLi-ASc and conventional AS parameters (AV V_max_ and mPG) were evaluated using scatterplots with Pearson correlation coefficients (PCC). To assess the predictive performance of baseline DLi-ASc for AS progression rate, we stratified patients by baseline DLi-ASc categories (<25, 25 to <50, 50 to <75, and ≥75) and compared the annualized progression rate of V_max_ and mPG across groups. These trends were visualized using violin plots and tested for significance using linear trend analysis. In addition, the associations between the estimated annualized progression rate of DLi-ASc, V_max_, and mPG – each estimated using LMMs - were also evaluated using scatterplots with PCC.

We assessed the predictive performance of both baseline DLi-ASc and its annualized rate of change for progression to severe AS. The earliest TTE demonstrating severe AS was designated as the event date, while patients who did not progress to severe AS were censored at their last available TTE. A competing risk analysis was performed using a Fine and Gray subdistribution hazard model.^14^ Two types of competing events were considered: (1) death occurring before the patient fulfilled echocardiographic criteria for severe AS, and (2) AV replacement (AVR) performed for other indications prior to meeting those criteria. The proportional hazards assumption was tested and confirmed to be satisfied.

Spline curve analyses were first performed to visualize the continuous association between each index (baseline value or annualized rate) and the subdistribution hazard of severe AS progression at 3 and 5 years, thereby identifying empirical inflection points. Based on these findings, baseline DLi-ASc was then evaluated both as a continuous variable (per 10-point increase) and as a categorical variable (<50, 50 to <60, 60 to <70, and ≥70). The annualized rate of change in DLi-ASc was assessed as a continuous variable (per 1-point/year increase) and in predefined categories (<1.0, 1.0–<1.2, 1.2–<1.4, and ≥1.4 points/year).

For both indices, we constructed Fine-Gray models under three adjustment strategies: (1) clinical factors only (age, sex, and estimated glomerular filtration rate [eGFR]), (2) clinical factors plus AV V_max_, and (3) clinical factors plus AV mPG. For models evaluating the annualized rate of change in DLi-ASc, baseline DLi-ASc was additionally included as a covariate. Variance inflation factors (VIF) were calculated for all models to assess collinearity. Subdistribution hazard ratios (HRs) with 95% confidence intervals (CIs) were reported for both continuous and categorical analyses. In addition, to exclude pre-diagnostic variability and validate the predictive performance of DLi-ASc and its annualized rate of change in a clinically confirmed AS population, we conducted a sensitivity analysis restricted to patients whose baseline TTE corresponded to the first examination at which AS was clinically diagnosed. The Fine and Gray competing risk model was reapplied in this subset using the same adjustment strategies to ensure consistency with the primary analysis.

All statistical analyses were performed using R software (v4.5.0; R Development Core Team, Vienna, Austria). Two-sided p-values <0.05 were considered statistically significant.

### 2.5. Funding

The study was supported by a grant from the Institute of Information & communications Technology Planning & Evaluation (IITP) funded by the Korea government (Ministry of Science and ICT); and the Medical AI Clinic Program through the NIPA funded by the MSIT. The funders had no role in study design, data collection and analysis, decision to publish, or preparation of the manuscript.

## 3. RESULTS

### 3.1. Baseline Characteristics

A total of 2,373 patients with 7,371 TTE examinations were included in the full analysis cohort. The training-excluded cohort, consisting of patients with no overlap with the DLi-ASc training dataset, comprised 1,117 patients and 3,376 TTEs (**Figure 1**). The median age was 76 years (interquartile range [IQR] 69–82) in the full cohort and 75 years (IQR 68-81) in the training-excluded cohort, with a similar sex distribution (**Table 1).** Baseline AS severity in the full cohort included normal AV (8.4%), AV sclerosis (18.9%), mild AS (53.2%), and moderate AS (19.4%). In the training-excluded cohort, the distribution shifted toward earlier disease stages, with a higher proportion of normal AV (17.6%) and AV sclerosis (26.9%), and a lower proportion of mild AS (34.6%). Median baseline DLi-ASc values were similar between the two groups: 49 (IQR 46–52) in the full cohort and 49 (IQR 45–52) in the training-excluded cohort. Median follow-up duration was slightly longer in the training-excluded cohort (47.8 months vs. 42.8 months), though the median number of follow-up TTEs was comparable (3 [IQR 2–4] vs. 3 [IQR 2–3]). Comparative baseline characteristics stratified by institution (SNUBH vs. CNUH) are presented in **Supplemental Results 1**.

**Table 1.** Baseline Characteristics of the Full Cohort and the Training-Excluded Cohort.

|  | Full Cohort<br>(n=2,373) | Training-Excluded Cohort<br>(n=1,117) |
| --- | --- | --- |
| Age | 76 (69–82) | 75 (68–81) |
| Male | 1,093 (46.1) | 523 (46.8) |
| Female | 1,280 (53.9) | 594 (53.2) |
| BSA | 1.7 (1.5–1.8) | 1.6 (1.5–1.8) |
| Baseline AS Severity |  |  |
| Normal | 200 (8.4) | 197 (17.6) |
| AV Sclerosis | 449 (18.9) | 301 (26.9) |
| Mild AS | 1,263 (53.2) | 386 (34.6) |
| Moderate AS | 461 (19.4) | 233 (20.9) |
| Baseline DLi-ASc | 49 (46–52) | 49 (45–52) |
| Follow-up, months | 42.8 (22.2–75.7) | 47.8 (21.0–87.8) |
| No. of TTEs | 7,371 | 3,376 |
| No. of TTE FU | 3 (2–4) | 3 (2–3) |
| At least 3 exams | 1,259 (53.1) | 569 (50.9) |
| At least 4 exams | 639 (26.9) | 276 (24.7) |

Values are given as numbers (percentage) or median (interquartile range).
Abbreviations: AS, aortic valve stenosis; AV, aortic valve; BSA, body surface area; CNUH, Chungnam National University Hospital; DLi-ASc, deep learning-derived index for the AS continuum; FU, follow-up

### 3.2. Temporal Changes in AS Severity and Corresponding DLi-ASc Distributions

The median follow-up duration from baseline TTE was 42.8 months (IQR 22.2–75.7 months) in the full cohort and 47.8 months (IQR 21.0-87.8 months) in the training-excluded cohort. As shown in **Figure 2**, the distribution of AS stages demonstrated a progressive shift toward more advanced stages over time in both cohorts, with increasing proportions of moderate and severe AS as the follow-up period extended (p for trend <0.001, based on cumulative link mixed model). This trend was consistently observed across all baseline AS severity categories (all p for trend < 0.001; **Supplemental Results 2**). Longitudinal trends of conventional AS parameters were examined using LMM. Annualized progression rates were 0.06 m/s/year (95% CI, 0.05–0.10) for V_max_ and 0.67 mmHg/year (95% CI, 0.34–1.34) for mPG in the full cohort, and 0.07 m/s/year (95% CI, 0.05–0.10) and 0.83 mmHg/year (95% CI, 0.50–1.44), respectively, in the training-excluded cohort (**Supplemental Results 3**).

**Figure 2.**
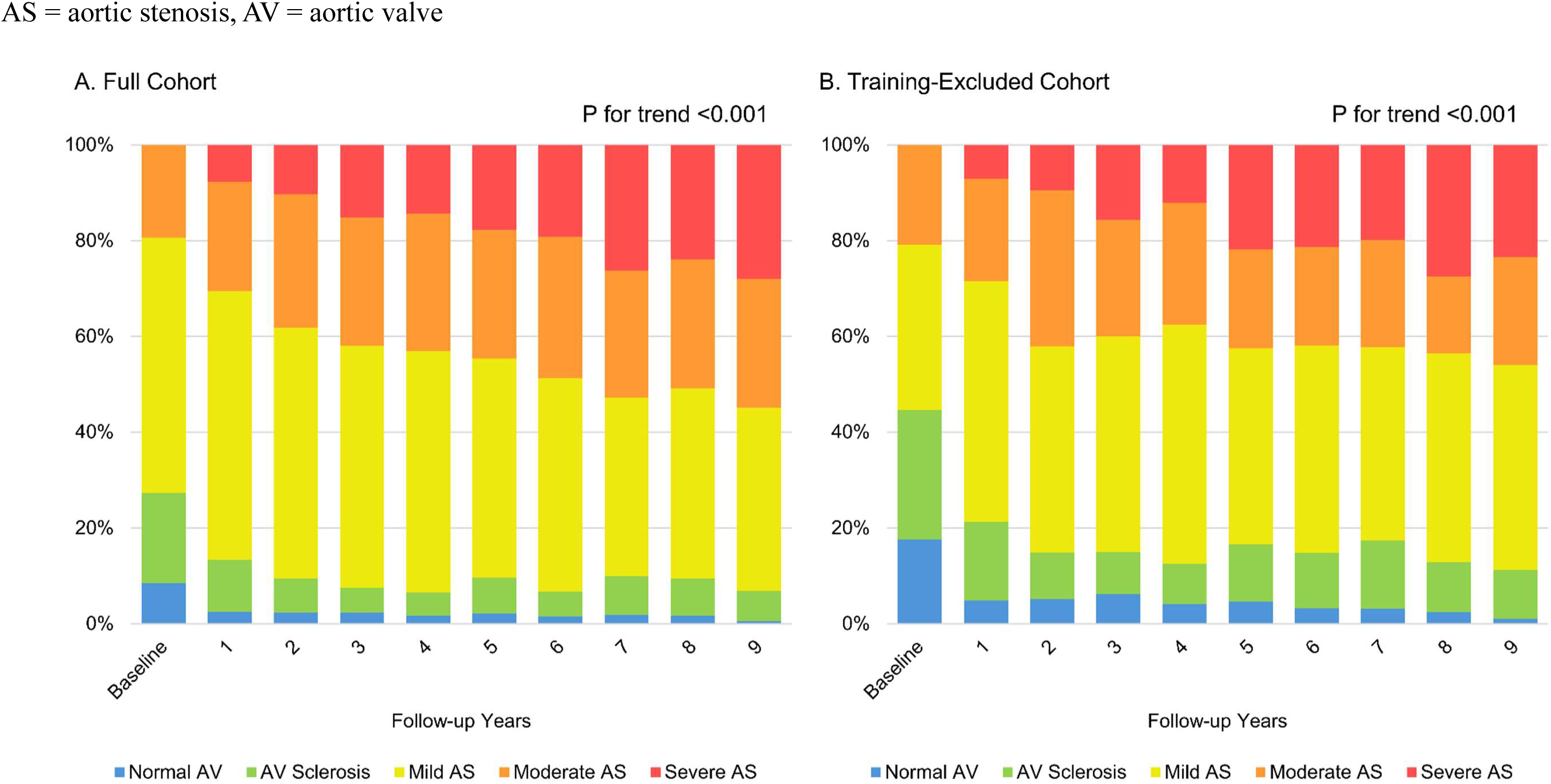
Temporal Progression of AS Severity Over Follow-Up Years.

In parallel, DLi-ASc also increased longitudinally across all baseline AS severity categories. As illustrated in **Figure 3**, the density distributions of DLi-ASc shifted progressively toward higher values over follow-up years, with all trends statistically significant (p for trend <0.001). Corresponding median DLi-ASc values at baseline and follow-up are summarized in **Supplemental Results 4**. For example, in the full cohort, DLi-ASc increased from 44 to 49 in normal AV, from 48 to 50 in AV sclerosis, from 49 to 53 in mild AS, and from 56 to 63 in moderate AS over 6 years. These patterns were also observed in the training-excluded cohort. The annualized rate of DLi-ASc progression was 1.11 (95% CI, 1.03-1.55) in the full cohort and 1.11 (95% CI, 1.02-1.23) in the training-excluded cohort, with progressively higher rates observed across worsening baseline severity categories (all p for trend <0.001; **Supplemental Results 3**).

**Figure 3.**
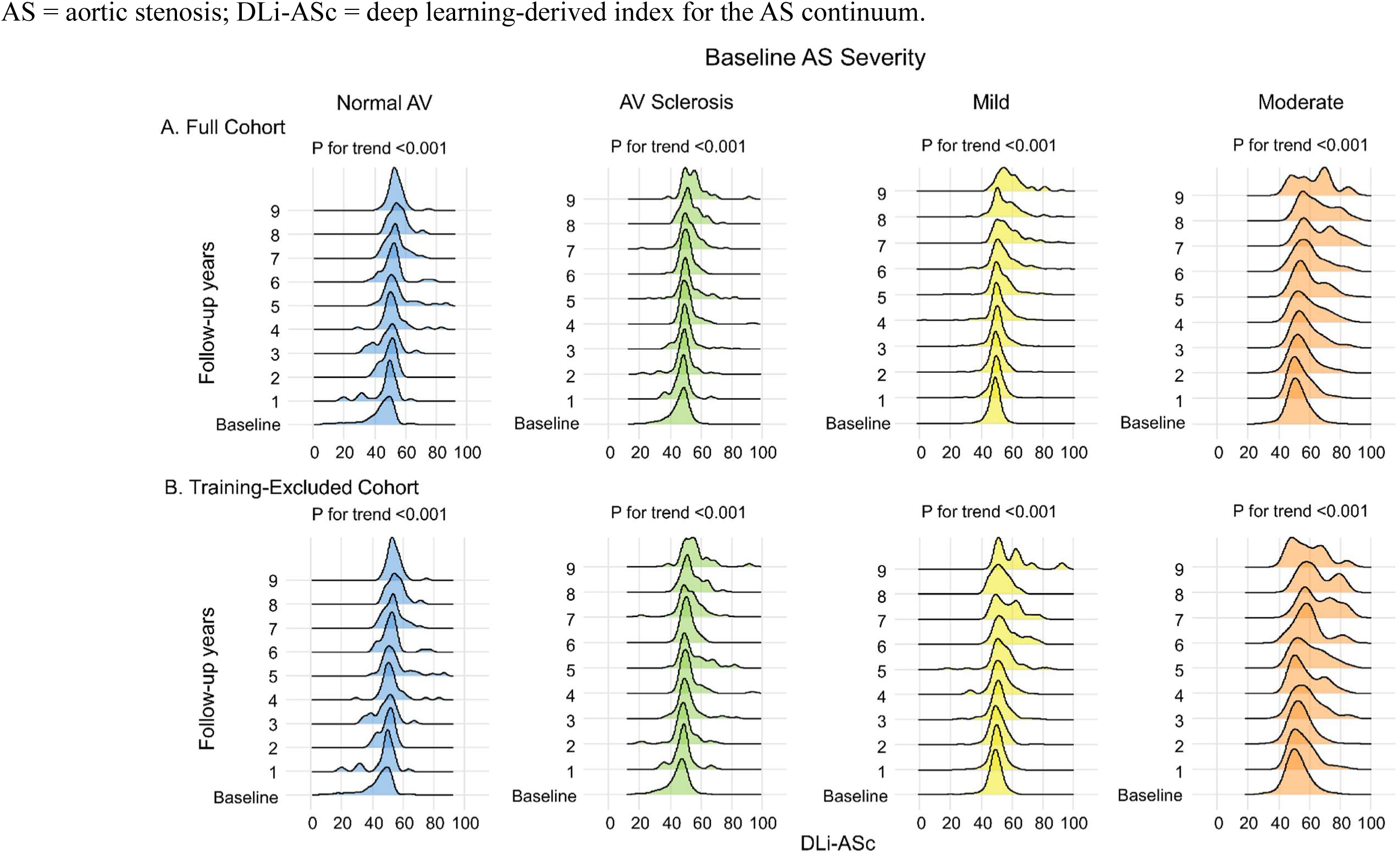
Temporal Trends in DLi-ASc Across Baseline AS Severity Categories.

For complementary visualization of individual-level DLi-ASc trajectories, subject-level connected line plots are also provided (**Supplemental Results 5**). These findings align with **Figure 3**, reinforcing DLi-ASc’s potential as a surrogate marker for monitoring long-term AS progression. While DLi-ASc generally increased over time, transient decreases were observed in 33.9% of consecutive TTE pairs, mostly when the follow-up interval was short (median 13 months). When comparing only the first and last TTEs (median 43 months), the proportion showing a net decrease dropped to 23.2%, again predominantly in cases with shorter follow-up. Notably, the absolute magnitude of DLi-ASc change increased with longer interscan intervals (**Supplemental Results 6**), suggesting that short-term decreases are more likely due to technical or image-related variability than true biological improvement.

### 3.3 Association of DLi-ASc with AS Severity and Progression

Analysis of all TTE examinations during follow-up revealed a significant positive correlation between DLi-ASc and conventional AS parameters in both the full and training-excluded cohorts, indicating that higher DLi-ASc values reflect greater hemodynamic severity. In the full cohort, PCC was 0.69 (95% CI: 0.67–0.70, p<0.001) for AV V_max_ and 0.66 (95% CI: 0.65-0.68, p<0.001) for AV mPG; in the training-excluded cohort, the corresponding values were 0.67 (95% CI: 0.65-0.69, p<0.001) and 0.61 (95% CI: 0.59-0.63, p<0.001), respectively (**Figure 4**). In addition, DLi-ASc also demonstrated significant correlation with AVA, though to a lesser degree than with V_max_ or mPG (**Supplemental Results 7**). PCC values were 0.59 (95% CI: 0.57-0.62, p<0.001) in the full cohort and 0.54 (95% CI: 0.47-0.60, p<0.001) in the training-excluded cohort.

**Figure 4.**
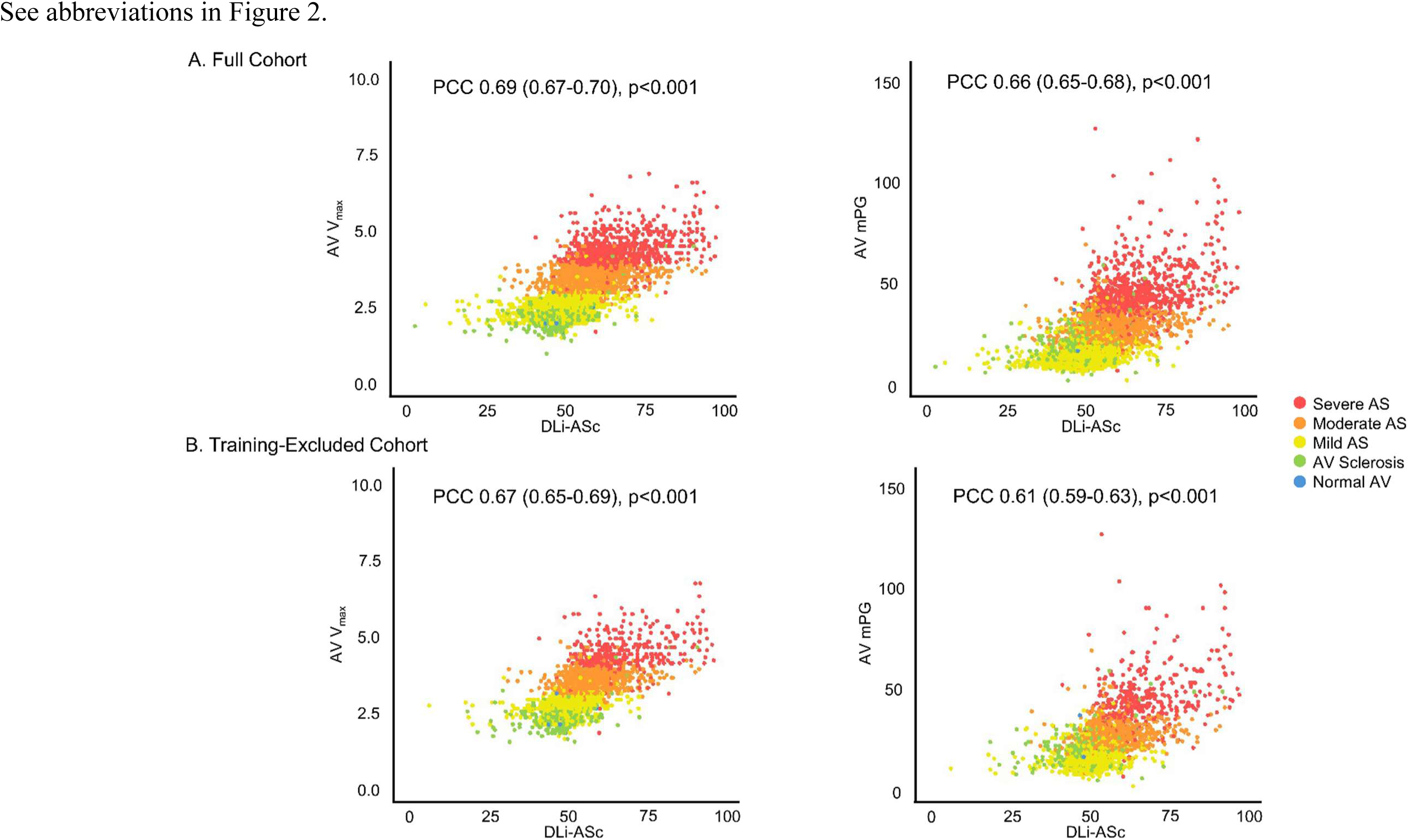
Association of DLi-ASc with Conventional Hemodynamic Parameters of AS.

Furthermore, stratification by baseline DLi-ASc categories revealed significant positive trends, with higher baseline DLi-ASc associated with faster progression of both AV V_max_ and mPG (all p for trend <0.001) (**Figure 5**). In addition, annualized progression parameters derived from LMMs (**Figure 6**) showed strong positive correlations between changes in DLi-ASc and corresponding changes in AV V_max_ (PCC = 0.71, 95% CI: 0.69–0.73, p < 0.001) and mPG (PCC = 0.68, 95% CI: 0.66–0.70, p < 0.001). The training-excluded cohort demonstrated consistent results, with PCC of 0.72 (95% CI: 0.69-0.74, p<0.001) for V_max_ and 0.66 (95% CI: 0.63-0.69, p<0.001) for mPG. These findings suggest that both the baseline level and longitudinal increase in DLi-ASc are closely aligned with the hemodynamic progression of AS.

**Figure 5.**
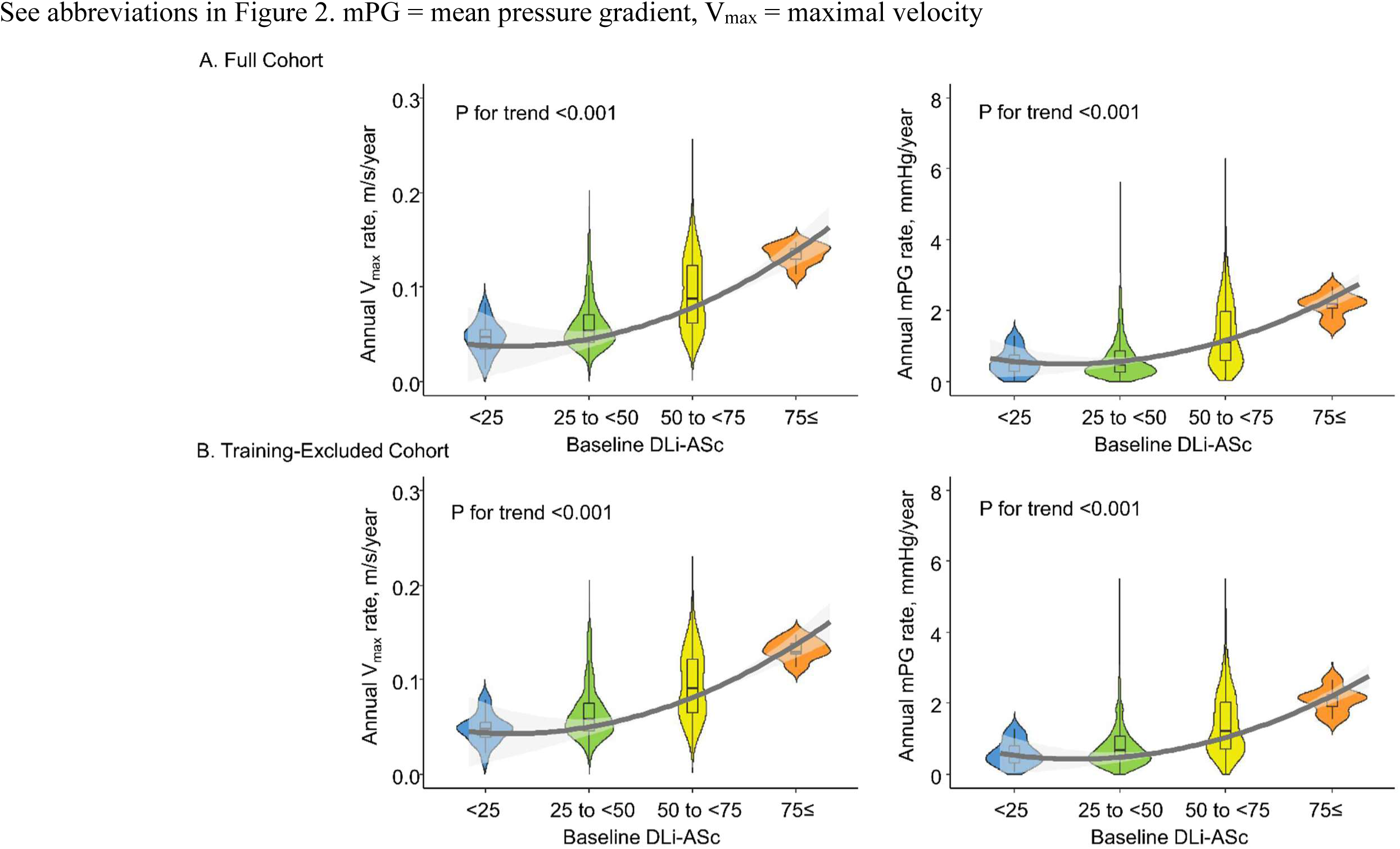
Baseline DLi-ASc and Its Association with Annualized Changes in AV V_max_ and mPG.

**Figure 6.**
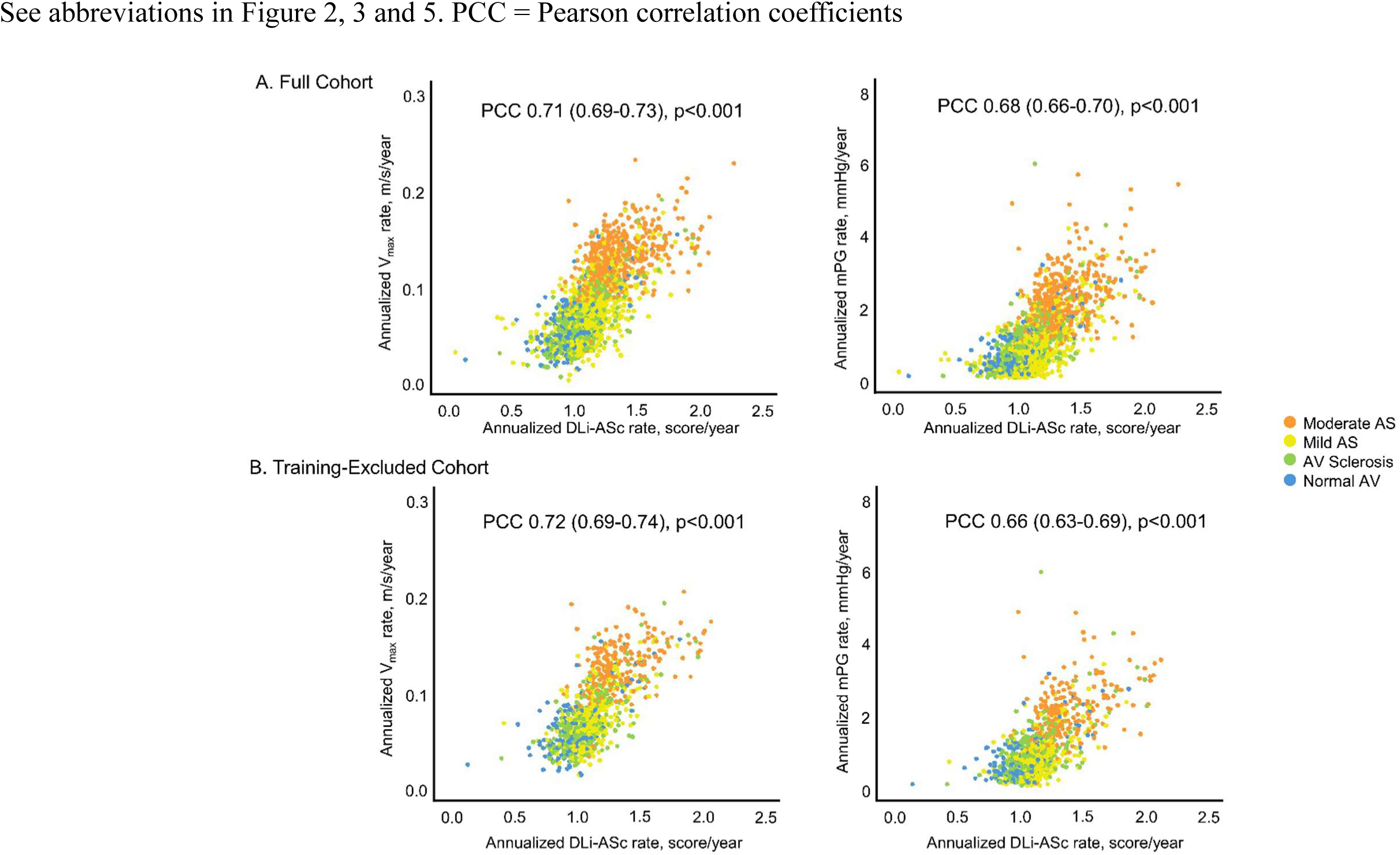
Association between Annual rates of DLi-ASc and Conventional AS Parameters.

### 3.4. Predictive Performance of DLi-ASc of Severe AS Progression

Spline curve analyses using Fine-Gray competing risk models demonstrated that the subdistribution hazard of severe AS progression at 3 and 5 years rose sharply once baseline DLi-ASc exceeded approximately 50, providing an empirical inflection point (**Supplemental Results 8**). Based on this observation, patients were stratified into predefined categories (<50, 50 to <60, 50 to <60, and *≥*70).

To further evaluate the independent prognostic value of DLi-ASc, we fitted competing risk model using three adjustment strategies: (1) clinical factors only (age, sex, and eGFR), (2) clinical factors plus AV V_max_, (3) clinical factors plus AV mPG. In the full cohort, baseline DLi-ASc remained a significant predictor under all models (**Table 2**). The adjusted HR per 10-point increase was 4.03 (95% CI: 3.40-4.78) with clinical factors only, 2.41 (95% CI: 2.09-2.78) after additional adjustment for AV V_max_, and 2.81 (95% CI: 2.41-3.29) after adjustment for mPG. Categorical DLi-ASc groups were also significantly associated with increased risk compared to the <50 reference group. For example, in the V_max_-adjusted model, DLi-ASc of 60–70 and ≥70 were associated with HRs of 4.59 (95% CI: 2.37–8.89) and 10.80 (95% CI: 5.60–20.81), respectively. In the training-excluded cohort, the adjusted HR per 10-point increase in DLi-ASc was 3.21 (95% CI: 2.65–3.89) with clinical factors only, 2.19 (95% CI: 1.82–2.62) with V_max_ adjustment, and 2.53 (95% CI: 2.08–3.07) with mPG adjustment. Consistent findings were observed for categorical DLi-ASc groups. VIFs for DLi-ASc and Doppler covariates were all <2.0 across models, indicating that multicollinearity was not a material concern.

**Table 2.** Prognostic Value of Baseline DLi-ASc for Severe AS Progression Under Different Adjustment Strategies.

|  | Unadjusted HR (95% CI) | Adjusted HR (95% CI)<br>(Clinical factors) | Adjusted HR (95% CI)<br>(Clinical factors<br>+ AV Vmax) | Adjusted HR (95% CI)<br>(Clinical factors<br>+ AV mPG) |
| --- | --- | --- | --- | --- |
| ***Full Cohort*** |  |  |  |  |
| DLi-ASc, per 10-score increase | 3.88 (3.18-4.74) | 4.01 (3.38-4.75) | 2.38 (2.07-2.76) | 2.80 (2.39-3.27) |
| DLi-ASc <50 | reference | reference | reference | reference |
| DLi-ASc 50 to <60 | 4.68 (3.81-5.74) | 4.86 (3.94-6.00) | 3.11 (2.55-3.80) | 3.66 (2.98-4.51) |
| DLi-ASc 60 to <70 | 24.23 (17.28-33.97) | 25.28 (17.96-35.59) | 4.53 (2.34-8.77) | 7.01 (3.84-12.78) |
| DLi-ASc 70 ≤ | 31.82 (15.12-66.93) | 33.18 (16.31-67.51) | 10.63 (5.51-20.50) | 16.13 (8.36-31.12) |
| ***Training-Excluded Cohort*** |  |  |  |  |
| DLi-ASc, per 10-score increase | 3.02 (2.40-3.80) | 3.16 (2.61-3.83) | 2.15 (1.80-2.58) | 2.49 (2.05-3.03) |
| DLi-ASc <50 | reference | reference | reference | reference |
| DLi-ASc 50 to <60 | 4.73 (3.56-6.29) | 4.85 (3.62-6.52) | 3.03 (2.30-3.99) | 3.54 (2.66-4.73) |
| DLi-ASc 60 to <70 | 15.53 (9.79-24.64) | 16.06 (10.08-25.57) | 3.50 (1.32-9.28) | 6.04 (2.52-14.47) |
| DLi-ASc 70 ≤ | 19.38 (5.89-63.77) | 20.13 (6.41-63.20) | 9.38 (3.47-25.36) | 13.82 (4.90-38.99) |

Clinical factors include age, sex, and estimated glomerular filtration rate (eGFR). Subdistribution HRs were estimated using Fine and Gray competing risk models for severe AS progression.
Abbreviations: AS, aortic valve stenosis; AV, aortic valve; CI, confidence interval; DLi-ASc, deep learning-derived index for the AS continuum; HR, hazard ratio; mPG, mean pressure gradient; Vmax, maximal transvalvular velocity.

Cumulative incidence curves accounting for death and AVR as a competing risk demonstrated a gradual increase in the risk severe AS progression across higher baseline DLi-ASc categories in both full and training-excluded cohorts (**Figure 7**). Notably, patients with baseline DLi-ASc ≥ 60 consistently showed greater cumulative incidence than those with lower DLi-ASc, the curves for the 60-70 and ≥70 groups appeared largely comparable over time.

**Figure 7.**
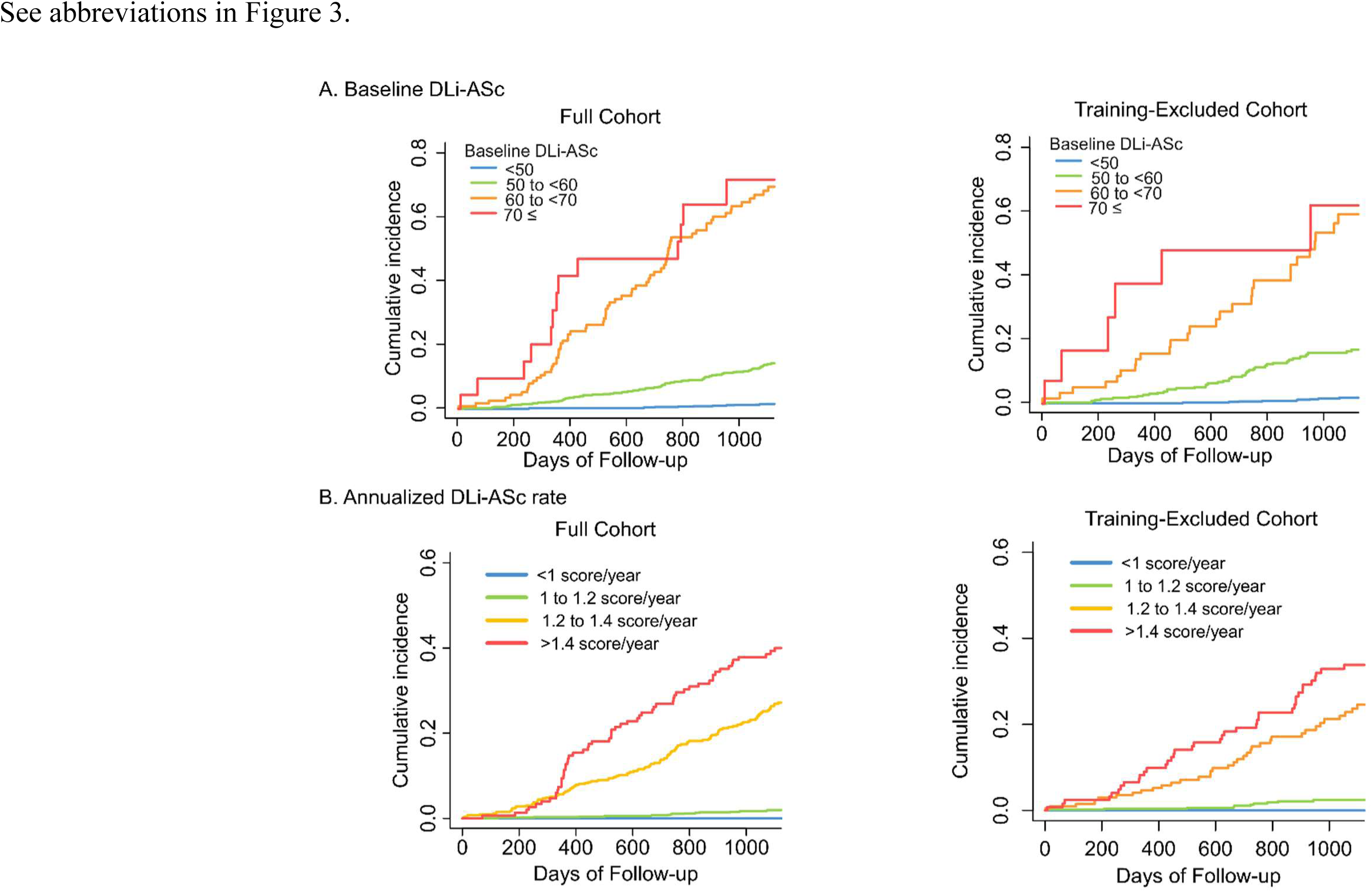
Cumulative Incidence of Severe AS Progression Stratified by Baseline DLi-ASc and Its Annualized Change.

To validate the predictive performance of DLi-ASc in a clinically confirmed AS population, we performed a sensitivity analysis excluding pre-diagnosis TTEs, defining baseline as the first exam at which AS was diagnosed (**Supplemental Results 9**). In this subset, DLi-ASc remained independently associated with severe AS progression across all adjustment models. For example, in the full cohort, the adjusted HR per 10-point increase was 1.86 (95% CI: 1.59–2.17) after AV V_max_ adjustment and 2.11 (95% CI: 1.79–2.49) after AV mPG adjustment. Similar associations were observed in the training-excluded cohort. These findings confirm the robustness of DLi-ASc as a prognostic marker even when restricted to post-diagnosis follow-up.

### 3.5. Prognostic Value of Annualized Rate of Change in DLi-ASc

Spline curve analyses demonstrated a generally linear increase in the subdistribution hazard of severe AS progression at 3 and 5 years with higher annualized rates of change in DLi-ASc, indicating a graded relationship between the trajectory of DLi-ASc and progression risk (**Supplemental Results 10**). To complement continuous analysis, we also evaluated categorical annualized rate group (<1.0, 1.0–<1.2, 1.2–<1.4, and ≥1.4 points/year).

We evaluated prognostic performance using three adjustment strategies, each including baseline DLi-ASc: (1) DLi-ASc plus clinical factors only (age, sex, and eGFR); (2) DLi-ASc plus clinical factors and baseline AV V_max_; and (3) DLi-ASc plus clinical factors and baseline AV mPG. The annualized rate remained an independent predictor across all models (**Table 3)**. When analyzed as a categorical variable, compared with <1.0 score/year, an increase ≥1.4 scores/year was associated with adjusted HRs of 12.30 (95% CI: 5.83-25.96) in the Vmax-adjusted model and 20.02 (95% CI: 9.17-43.71) in the mPG-adjusted model. VIFs for baseline DLi-ASc, its annualized rate, and Doppler covariates were all <2.5 across models, indicating no evidence of problematic multicollinearity.

**Table 3.** Prognostic Value of Annualized Rate of Change in DLi-ASc for Severe AS Progression Under Different Adjustment Strategies.

|  | Adjusted HR (95% CI)<br>(Baseline DLi-ASc) | Adjusted HR (95% CI)<br>(Clinical factors +<br>Baseline DLi-ASc) | Adjusted HR (95% CI)<br>(Clinical factors<br>+ AV Vmax + Baseline DLi-ASc) | Adjusted HR (95% CI)<br>(Clinical factors<br>+ AV mPG + Baseline<br>DLi-ASc) |
| --- | --- | --- | --- | --- |
| ***Full Cohort*** |  |  |  |  |
| Annualized DLi-ASc increasing<br>rate, per score/year | 13.25 (8.87-19.79) | 13.07 (8.84-19.32) | 2.39 (1.19-4.80) | 3.88 (2.03-7.42) |
| <1 score/year | reference | reference | reference | reference |
| 1 to 1.2 score/year | 3.26 (3.08-12.7) | 6.54 (3.21-13.33) | 5.58 (2.76-11.25) | 6.38 (3.13-12.99) |
| 1.2 to 1.4 score/year | 32.15 (15.90-65.00) | 33.03 (16.32-66.86) | 17.20 (8.12-36.45) | 23.32 (11.15-48.78) |
| >1.4 score/year | 42.51 (20.94-86.30) | 43.65 (21.51-88.57) | 13.60 (6.20-29.84) | 20.02 (9.17-43.71) |
| ***Training-Excluded Cohort*** |  |  |  |  |
| Annualized DLi-ASc increasing<br>rate, per score/year | 15.52 (10.03-24.00) | 15.25 (9.92-23.44) | 2.22 (1.29-3.83) | 3.63 (2.14-6.14) |
| <1 score/year | reference | reference | reference | reference |
| 1 to 1.2 score/year | 5.38 (2.49-11.58) | 5.48 (2.54-11.84) | 4.72 (2.24-9.94) | 5.32 (2.51-11.28) |
| 1.2 to 1.4 score/year | 25.50 (11.82-55.02) | 25.30 (11.72-54.60) | 13.25 (5.95-29.50) | 17.40 (8.08-37.46) |
| >1.4 score/year | 36.56 (17.14-78.02) | 36.54 (17.11-78.06) | 11.08 (4.75-25.86) | 15.33 (6.93-33.94) |

Clinical factors include age, sex, and estimated glomerular filtration rate (eGFR). Subdistribution HRs were estimated using Fine and Gray competing risk models for severe AS progression.
Abbreviations: AS, aortic stenosis; AV, aortic valve; CI, confidence interval; DLi-ASc, deep learning-derived index for the AS continuum; HR, hazard ratio; mPG, mean pressure gradient; Vmax, maximal transvalvular velocity.

Cumulative incidence analyses further demonstrated that patients with a rapid annualized increase (≥1.2 points/year) had a substantially higher risk of progression to severe AS compared with those with slower progression (**Figure 7**). These findings highlight that both baseline DLi-ASc and its trajectory provide complementary prognostic information, supporting a more dynamic approach to patient surveillance.

All analyses were repeated in the training-excluded cohort (**Table 3**, **Figure 7**) and in a subset excluding pre-diagnosis TTEs (**Supplemental Results 11**), and results were consistent across both sensitivity analyses, confirming the robustness of these findings.

## 4. DISCUSSION

In this longitudinal study, we applied previously validated DLi-ASc to serial TTE examination. Both baseline score and annualized rate of change were strongly associated with progression of conventional echocardiographic parameters (AV V_max_ and mPG) and independently predicted severe AS progression in competing risk-adjusted models. By quantifying temporal score change as annualized rates, we demonstrated that DLi-ASc not only mirrors the natural trajectory of AS but also provides prognostic information beyond baseline risk stratification. Taken together, these findings highlight the complementary value of baseline DLi-ASc and its trajectory: the baseline score identifies patients predisposed to faster progression, while the rate of change refines risk estimation during follow-up and enables more dynamic disease monitoring.

The role of AI in TTE is evolving. Early applications focused on automating manual measurements, enhancing accuracy, and reproducibility. With most TTE parameters now capable of automation, AI has expanded to disease severity assessment, as seen in diastolic dysfunction^15,16^ and AS evaluation.^8^ More recently, AI has progressed beyond parameter-based analysis to DL-based direct TTE video analysis, aiming to mimic expert visual assessment. Some efforts have targeted specific conditions with high clinical relevance, such as AS,^5–7^ diastolic dysfunction,^15^ hypertrophic cardiomyopathy, and pericardial disease,^17,18^ developing task-oriented models that provide precise evaluation of a single disease. In parallel, similar research efforts have also pursued comprehensive multitask or foundation models – such as PanEcho,^19^ EchoPrime,^20^ EchoClip,^21^ and EchoFM^22^ – that aim to replicate the global synthesis performed by expert echocardiographers across multiple findings. In our previous study, we developed a DL model capable of accurately assessing AS severity using PLAX and/or PSAX views.^5^ The strength of this DL-based index lies in its ability to mimic visual analysis while providing a quantitative digital marker. Our prior work validated this approach in a large cross-sectional cohort, confirming its diagnostic performance. However, while the model was designed to reflect the AS disease continuum, whether DLi-ASc would increase in parallel with AS progression over time remained unclear.

Several DL-based indices have been proposed for AS severity assessment,^5–7^ differing in input views, labeling strategies, and intended clinical applications. Holste et al. introduced a PLAX view-based model trained to detect severe AS (Digital AS Severity Index, DASSi) and demonstrated high diagnostic performance in both internal and external test cohorts.^6^ A subsequent follow-up study by Oikonomou et al. extended the evaluation of this same model to baseline prognostic analyses and longitudinal follow-up, showing that baseline scores predicted subsequent severe AS and AVR.^9^ However, their longitudinal analysis did not directly assess whether the DL index changed in parallel with AS progression. In contrast, our analysis is the first to demonstrate that DLi-ASc progressively increased over serial follow-up, closely mirroring the natural course of AS. Furthermore, by quantifying temporal changes as annualized rates, we showed that these trajectories tracked with concurrent changes in conventional hemodynamic parameters and independently predicted severe AS progression, even after adjustment for baseline DLi-ASc and Doppler indices. This extends the role of DL-based indices beyond baseline prognostication, positioning DLi-ASc as a tool for both static and dynamic risk assessment.

Cumulative incidence analyses, together with categorical Fine-Gray models, consistently demonstrated that both higher baseline scores (particularly ≥60) and rapid annualized increases (≥1.2 points/year) were associated with sharp rise in the risk of severe AS progression. This concordance across analytic approaches reinforces the robustness of these thresholds and underscores the complementary value of baseline level and longitudinal trajectory in identifying patients at higher risk. These findings support the potential role of DLi-ASc in guiding personalized surveillance strategies: rather than applying uniform monitoring intervals to all AS patients, integrating both baseline and longitudinal information may enable earlier detection of rapid progression and tailoring follow-up intensity. In addition, although current AS management primarily focuses on AV replacement after severe AS develops, the ability to quantify AS progression with DLi-ASc could become particularly valuable if medical treatment to slow AS progression becomes available in the future. In such contexts, both baseline and longitudinal DLi-ASc could serve as objective digital markers for monitoring treatment response and identifying patients who may benefit from earlier or more aggressive intervention. Despite its promise, clinical implementation of AI-based tools like DLi-ASc may be influenced by factors such as accessibility, cost, and integration into existing echocardiographic workflows. Further research should validate its clinical applicability across diverse settings and to develop strategies for seamless implementation in standardized clinical pathways.

This study has several limitations. First, the retrospective nature of data collection introduces inherent constraints. In particular, the dataset was derived from electronic health records (EHRs), where TTEs were performed based on clinical indications rather than a standardized protocol. This indication-driven acquisition may introduce variability in follow-up timing and disease spectrum, which could affect the generalizability of our findings, especially when extending validation to community-based cohorts. Irregular TTE intervals, inherent to such indication-driven follow-up, limited the precision of annual progression estimates based on either conventional AS parameters or DLi-ASc. To address this, we employed LMMs, which account for repeated measures and variable follow-up intervals, thereby reducing bias from irregular TTE schedules and improving the robustness of the estimated progression rate. While a prospective study with a standardized follow-up protocol would be the most rigorous design for validating DLi-ASc as a longitudinal monitoring tool, such studies require substantial time, resources, and patient enrollment. Large-scale retrospective analyses such as the present work are, therefore, a necessary first step to generate the preliminary evidence needed to justify and design such prospective investigations. Moreover, most real-world AS surveillance occurs in non-protocolized environments; thus, our findings provide insight into the potential performance of DLi-ASc in the setting where it would most likely be applied.

In addition, although commonly used,^9,23^ progression tracking relied primarily on flow-dependent metrics such as V_max_ and mPG, which are susceptible to variations in stroke volume and hemodynamic loading conditions. However, diagnosis of severe AS was not based solely on these parameters, but rather on comprehensive clinical interpretation that incorporated flow status documented in TTE reports and, when available, results from additional diagnostic tests. For future validation, prospective studies with standardized follow-up protocols – incorporating both flow-dependent and flow-independent measures such as CT-based AV calcium scoring – could further validate the value of DLi-ASc in assessing the temporal dynamics of AS progression in a more systematic manner and may also enable advanced modelling approaches – such as trajectory clustering or unsupervised pattern recognition – to explore distinct AS progression phenotypes.

Furthermore, although coronary artery disease (CAD) is a major competing risk in patients with mild to moderate AS, CAD-specific diagnostic or outcome data were not uniformly available due to the retrospective design. To mitigate potential bias, we applied a Fine and Gray competing risk model, treating death as a competing event. Still, the lack of adjudicated CAD outcomes remains a limitation, and future studies should incorporate comprehensive ischemic event data to further evaluate the prognostic value of DLi-ASc.

The dataset was derived from two tertiary hospitals in South Korea, where TTE examinations were performed by specialized cardiologists with expertise in echocardiography. While this ensures high-quality imaging and reliable data, it also raises concerns about generalizability to resource-limited settings, where non-experts often perform TTE. Since DLi-ASc was developed to enhance AS assessment across diverse clinical environments, further studies are needed to evaluate its performance in settings with limited echocardiographic expertise. In addition, the study cohort was restricted to patients who had AS diagnosed either at baseline or during follow-up, with exclusion of those with discordant AS classification or moderate-to-severe concomitant valvular disease. This strategy was intended to minimize confounding when tracking AS progression with flow-dependent parameters, but it inevitably narrows the study population and may limit broader applicability. Extending validation to a more inclusive population – particularly those without significant AS at baseline – remains an important direction for future research. Additionally, while DL analysis was successfully performed in all cases and DLi-ASc generally increased over time, transient decreases were occasionally observed, particularly with short follow-up intervals. These likely reflect technical or image-related variability, representing an inherent limitation of DL-based image analysis. Finally, to ensure broader generalizability, additional research in diverse racial and geographic populations is essential to establish the clinical applicability and real-world implications of DLi-ASc.

## 5. CONCLUSION

This study demonstrated the utility of DLi-ASc as a dynamic marker for longitudinal monitoring of AS and for predicting progression to severe AS. Both baseline levels and the annualized rate of change were significantly associated with faster hemodynamic progression and independently predicted future severe AS in competing risk-adjusted models. These findings underscore the complementary value of baseline and longitudinal trajectories, supporting the potential of DLi-ASc as a noninvasive imaging biomarker to track disease course, identify high-risk patients, and guide personalized surveillance strategies.

## Data Availability

Please contact the corresponding author to request the minimal anonymised dataset. Researchers with additional inquiries about the deep learning model developed in this study are also encouraged to reach out to the corresponding author.

## Contributors

All authors contributed equally to this study. All authors have read and approved the final version of the manuscript. J.P, J.K, and Y.E.Y verified the underlying data of the current study.

## Supplemental Materials

Supplemental Results 1. Baseline Characteristics by Institution

Supplemental Results 2. Longitudinal Trends in AS Severity Stratified by Baseline AS Severity

Supplemental Results 3. Annualized rate of AV V_max_, mPG, and DLi-ASc

Supplemental Results 4. The yearly trends in DLi-ASc distribution according to baseline AS severity

Supplemental Results 5. Longitudinal Trajectories of DLi-ASc by Baseline AS Severity

Supplemental Results 6. Short-term Fluctuations and Time-Dependent Increase in DLi-ASc During Follow-up.

Supplemental Results 7. Association of DLi-ASc with AVA

Supplemental Results 8. Spline Curves of Subdistribution Hazard for Severe AS Progression by Baseline DLi-ASc

Supplemental Results 9. Risk of Severe AS Progression Stratified by DLi-ASc at Time of Clinical Diagnosis

Supplemental Results 10. Spline Curves of Subdistribution Hazard for Severe AS Progression by Annualized Change in DLi-ASc

Supplemental Results 11. Risk of Severe AS Progression Stratified by Annualized Change in DLi-ASc at Time of Clinical Diagnosis

## REFERENCES

1 Owens DS, Bartz TM, Buzkova P, Massera D, Biggs ML, Carlson SD, et al. Cumulative burden of clinically significant aortic stenosis in community-dwelling older adults. Heart. 2021;107:1493–1502.

2 Yi B, Zeng W, Lv L, Hua P. Changing epidemiology of calcific aortic valve disease: 30-year trends of incidence, prevalence, and deaths across 204 countries and territories. Aging. 2021;13:12710–12732.

3 Otto CM, Nishimura RA, Bonow RO, Carabello BA, Erwin JP 3rd, Gentile F, et al. 2020 ACC/AHA Guideline for the Management of Patients With Valvular Heart Disease: A Report of the American College of Cardiology/American Heart Association Joint Committee on Clinical Practice Guidelines. J Am Coll Cardiol. 2021;77:e25–e197.

4 Baumgartner H, Hung J, Bermejo J, Chambers JB, Edvardsen T, Goldstein S, et al. Recommendations on the Echocardiographic Assessment of Aortic Valve Stenosis: A Focused Update from the European Association of Cardiovascular Imaging and the American Society of Echocardiography. J Am Soc Echocardiogr. 2017;30:372–392.

5 Park J, Kim J, Jeon J, Yoon YE, Jang Y, Jeong H, et al. Artificial intelligence-enhanced comprehensive assessment of the aortic valve stenosis continuum in echocardiography. EBioMedicine. 2025;112:105560.

6 Holste G, Oikonomou EK, Mortazavi BJ, Coppi A, Faridi KF, Miller EJ, et al. Severe aortic stenosis detection by deep learning applied to echocardiography. Eur Heart J. 2023;44:4592–4604.

7 Wessler BS, Huang Z, Long GM Jr, Pacifici S, Prashar N, Karmiy S, et al. Automated Detection of Aortic Stenosis Using Machine Learning. J Am Soc Echocardiogr. 2023;36:411–420.

8 Krishna H, Desai K, Slostad B, Bhayani S, Arnold JH, Ouwerkerk W, et al. Fully Automated Artificial Intelligence Assessment of Aortic Stenosis by Echocardiography. J Am Soc Echocardiogr. 2023;36:769–777.

9. Oikonomou EK, Holste G, Yuan N, Coppi A, McNamara RL, Haynes NA, et al. A Multimodal Video-Based AI Biomarker for Aortic Stenosis Development and Progression. JAMA Cardiol. 2024;9:534–544.

10. Oikonomou EK, Craig NJ, Holste G, Shankar SV, White A, Mahendran M, et al. Artificial intelligence-enabled echocardiography as a surrogate for multi-modality aortic stenosis imaging: post-hoc analysis of a clinical trial. medRxiv. 2025.03.26.25324690. doi: 10.1101/2025.03.26.25324690.

11 Collins GS, Reitsma JB, Altman DG, Moons KG. Transparent Reporting of a multivariable prediction model for Individual Prognosis or Diagnosis (TRIPOD): the TRIPOD statement. Ann Intern Med. 2015;162:55–63.

12 Christensen RHB. ordinal-regression models for ordianl data. R-package Version 2023.12-4. The R Foundation. 2023. Retrieved from https://CRAN.R-project.org/package=ordinal.

13 Bates D, Mächler M, Bolker B, Walker S. Fitting Linear Mixed-Effects Models Using lme4. Journal of Statistical Software. 2015;67:1–48.

14 Gray B. cmprsk: Subdistribution Analysis of Competing Risks. R package version 2.2-15 The R Foundation. 2024. Retrieved from https://CRAN.R-project.org/package=cmprsk.

15 Chen X, Yang F, Zhang P, Lin X, Wang W, Pu H, et al. Artificial Intelligence-Assisted Left Ventricular Diastolic Function Assessment and Grading: Multiview Versus Single View. J Am Soc Echocardiogr. 2023;36:1064–1078.

16 Park J, Jeon J, Yoon YE, Jang Y, Kim J, Jeong D, et al. Artificial intelligence-enhanced automation of left ventricular diastolic assessment: a pilot study for feasibility, diagnostic validation, and outcome prediction. Cardiovasc Diagn Ther. 2024;14:352–366.

17 Park J, Kim J, Jeon J, Yoon YY, Jang Y, Jeong H, et al. Single View Echocardiographic Analysis for LVOT Obstruction Prediction in Hypertrophic Cardiomyopathy: A Deep Learning Approach. J Am Soc Echocardiogr. In press 2025.

18 Jeong S, Moon IT, Jeon J, Jeong D, Lee J, Kim J, et al. Deep Learning-Based Multi-View Echocardiographic Framework for Comprehensive Diagnosis of Pericardial Disease. medRxiv. 2025.07.24.25332108. doi: 10.1101/2025.07.24.25332108.

19 Holste G, Oikonomou EK, Tokodi M, Kovács A, Wang Z, Khera R. Complete AI-Enabled Echocardiography Interpretation With Multitask Deep Learning. JAMA. 2025;334:306–318.

20 Vukadinovic M, Tang X, Yuan N, Cheng P, Li D, Cheng S, et al. EchoPrime: A multi-video view-informed vision-language model for comprehensive echocardiography interpretation. arXiv [cs.CV; 2024]. Available from: https://arxiv.org/abs/2410.09704

21 Christensen M, Vukadinovic M, Yuan N, Ouyang D. Vision-language foundation model for echocardiogram interpretation. Nat Med. 2024;30:1481–1488.

22 Kim S, Jin P, Song S, Chen C, Li Y, Ren H, et al. EchoFM: Foundation Model for Generalizable Echocardiogram Analysis. arXiv [cs.CV; 2024]. Available from: https://arxiv.org/abs/2410.23413

23 Willner N, Prosperi-Porta G, Lau L, Nam Fu AY, Boczar K, Poulin A, et al. Aortic Stenosis Progression: A Systematic Review and Meta-Analysis. JACC Cardiovasc Imaging. 2023;16:314–328.

